# Evaluating Allopregnanolone as a Potential Mediator of Prenatal Psychosocial Distress and Birth Outcomes in the Healthy Start Cohort

**DOI:** 10.64898/2026.07.26.26358958

**Authors:** Gabriella Mayne, K. Joseph Hurt, Sara Yeatman, Jost Klawitter, David P. Tracer, Uwe Christians, Dana Dabelea, Wei Perng

## Abstract

**Background:** Prenatal psychosocial distress is a risk factor for adverse birth outcomes such as preterm birth, but the biological pathways remain incompletely understood. Allopregnanolone (ALLO), a stress-responsive, progesterone-derived neuroactive steroid, may contribute to pregnancy maintenance by inhibiting uterine activation and modulating inflammatory and neuroendocrine pathways involved in parturition. Few human studies have examined ALLO during pregnancy in relation to birth timing and related outcomes, and no study to our knowledge has examined the entire pathway of prenatal psychosocial distress, ALLO, and birth outcomes. Here, we evaluated whether maternal ALLO concentrations and the ALLO-to-progesterone ratio mediate associations between prenatal psychosocial distress and birth outcomes.

**Methods:** This study included 237 pregnant participants from the Healthy Start Cohort, enriched for psychosocial distress (n=57 high-distress; n=180 low-distress) assessed using the Edinburgh Perinatal Depression Scale (EPDS) and EPDS-3A anxiety subscale. We measured maternal serum ALLO and related steroid hormones at approximately 17 and 27 weeks’ gestation (range: 10–34 weeks) using a validated HPLC-MS/MS assay and evaluated natural log (ln)-transformed ALLO and the ALLO-to-progesterone ratio as potential mediators. The primary outcome was gestational age at birth; secondary outcomes included birthweight-for-gestational-age z-score (BW/GA), percent fat mass (%FM), and birth length-for-gestational-age z-score (BL/GA). We conducted regression-based mediation analyses by comparing the total and direct effects of prenatal psychosocial distress after adjustment for each proposed mediator.

**Results:** Participants had a mean ± SD age of 29±6 years; 38% were nulliparous, and 60% identified as non-Hispanic White. Of the birth outcomes assessed, prenatal psychosocial distress was only associated with BL/GA (β = -0.49; 95% CI: -0.84, -0.14). Adjustment for ALLO at ∼27 weeks as a mediator minimally attenuated the association between prenatal distress and BL/GA (β = -0.46; 95% CI: -0.81, -0.10, 6.1% attenuation), whereas adjustment for the ALLO-to-progesterone ratio resulted in 20.8% attenuation (β = -0.38; 95% CI: -0.74, -0.03).

**Conclusions:** Maternal prenatal distress was associated with shorter birth length. The ALLO-to-progesterone ratio, but not ALLO itself, may partially mediate this relationship. These findings support neurosteroid metabolism as a biological pathway linking prenatal distress to fetal length accrual. Future mechanistic studies are needed to confirm these findings.

## INTRODUCTION

Prenatal psychosocial distress – commonly manifesting as depression and/or anxiety during pregnancy – is an established risk factor for adverse birth outcomes, including preterm birth, low birth weight, and impaired fetal growth [1–7]. Psychosocial distress may influence birth timing and fetal growth through overlapping neuroendocrine, inflammatory, vascular, and metabolic pathways involved in both the maternal stress response and pregnancy maintenance [8,9]. Despite decades of evidence linking maternal psychosocial distress with adverse birth outcomes, the biological mechanisms remain incompletely understood [9]. Identifying these mechanisms may help reveal clinically actionable targets for prevention and intervention.

Allopregnanolone (ALLO), a progesterone-derived neuroactive steroid, is a promising candidate mechanism. During acute stress, circulating ALLO concentrations increase as part of a homeostatic response [10]. In contrast, chronic stress states, including depression and anxiety during the perinatal period, have been associated with lower circulating ALLO concentrations [11–18], though the literature in humans is inconsistent [19–29]. The efficacy of synthetic ALLO formulations for postpartum depression highlights the clinical relevance of ALLO-responsive pathways [30–32].

ALLO also plays a key role in pregnancy physiology [33,34]. Animal and in vitro studies, suggest that ALLO may contribute to pregnancy maintenance by promoting uterine quiescence [35], suppressing inflammatory pathways [36], and regulating corticotropin-releasing hormone [37], a central mediator of parturition [38]. Consistent with this biology, pharmacologic inhibition of ALLO synthesis in late-pregnant rats shortened gestation, reduced pup viability, and impaired cognitive and neuroendocrine development [39].

Despite strong biological plausibility, evidence linking endogenous maternal ALLO concentrations with birth outcomes in humans remains limited. A nested case-control study found no difference in maternal serum ALLO in individuals who gave birth preterm birth versus gestational age matched controls that gave birth at term [40]. Similarly, Turkmen et al. observed no difference in maternal plasma ALLO concentrations in Swedish individuals admitted to the hospital with preterm labor symptoms in comparison to age-matched controls [41]. However, both studies were relatively small (n≤54) and neither evaluated gestational age at birth as a continuous outcome, potentially limiting the ability to detect smaller but biologically meaningful associations. Even shorter gestational duration within the term range have been associated with poorer offspring neurodevelopmental and cognitive outcomes [42–44]. To date, we are unaware of any published human study evaluating the complete pathway linking prenatal psychosocial distress, ALLO-related maternal neurosteroid biology, and birth outcomes. A prospective study addressing this question has been proposed but results are not yet available [45].

To address these gaps, we build upon our prior work in the Healthy Start Cohort in which high psychosocial distress was associated with 20% lower maternal ALLO concentrations in minimally adjusted models and a consistently lower ALLO-to-progesterone ratio across early (∼17 weeks’ gestation) and mid-pregnancy (∼27 weeks’ gestation) [18]. These findings suggest alterations in neurosteroid biosynthesis may represent physiological correlates of prenatal psychosocial distress and a potential mechanism through which maternal distress influences birth outcomes. In the present study, we evaluated whether maternal ALLO concentrations and the ALLO-to-progesterone ratio mediate associations between prenatal psychosocial distress and birth timing and neonatal anthropometry. We hypothesized that high prenatal psychosocial distress would be associated with shorter gestation and less favorable neonatal anthropometry as indicated by shorter birth length, lower birth weight, and differences in neonatal adiposity, and that maternal ALLO concentrations and the ALLO-to-progesterone ratio would partially mediate these associations.

## METHODS

### Study population

We used banked specimens from the Healthy Start cohort, a prospective pre-birth cohort of 1,410 pregnant individuals recruited at the University of Colorado Anschutz Medical Campus (Denver, CO, USA) between December 2009 and May 2014 [46]. All participants provided written informed consent, and the Colorado Multiple Institutional Review Board approved the parent study and this secondary analysis (COMIRB #09-0563 and #24-0224).

Participant selection and psychosocial distress classification have been described previously [18]. Briefly, all participants in Healthy Start Cohort had data on the primary outcome of interest, gestational age at birth. Therefore, we subsequently selected participants for this analysis using an extreme phenotyping approach based on Edinburgh Perinatal Depression Scale (EPDS) scores and the EPDS-3A anxiety subscale. Participants were classified as high distress if they had an EPDS score ≥13 and/or an EPDS-3A score ≥7 at either study visit, and as low distress if they had EPDS <4 and EPDS-3A <2 at both visits. This approach yielded an analytic sample of 237 participants (57 high distress and 180 low distress). Morning fasting blood samples and psychosocial questionnaires were collected at approximately 17 and 27 weeks’ gestation. Blood samples were processed immediately and stored at −80°C until analysis.

### Neurosteroid Measurement

We quantified maternal serum allopregnanolone (ALLO) and its key precursor hormone, progesterone, using a validated high-performance liquid chromatography–tandem mass spectrometry (HPLC–MS/MS) assay. Assay methods have been described previously [18,47]. The assay met established clinical and industry performance criteria [48]. The working range of the assay for ALLO was 0.39–100 ng/mL and for progesterone was 1.56–400 ng/mL.

For the present study, we considered ALLO concentrations measured at two time points during pregnancy (mean 17 and 27 weeks’ gestation, range 10–34 weeks), as well as change in ALLO across the two time points (ΔALLO = Time 2 – Time 1), and the ALLO-to-progesterone ratio as an indicator of neurosteroid metabolism of *a priori* interest based on our prior findings [18]. For regression analyses, ALLO and the ALLO-to-progesterone ratio were natural log(ln)-transformed to combat deviations from normality and z-score standardized for interpretability.

### Birth Outcomes

Study staff abstracted gestational age at birth (weeks) from the medical record, and measured birth length and weight within 72 hours of delivery. Additionally, study staff measured neonatal body composition using the PEA POD system (Life Measurement, USA), which uses air displacement plethysmography to estimate fat mass and fat-free mass from body weight and body volume. This method is safe, noninvasive, highly reproducible, and well validated against reference methods including dual-energy X-ray absorptiometry (DXA) [49].

The primary outcome of interest in this analysis was gestational age at birth assessed continuously (weeks) and dichotomized as preterm birth yes/no (<37 vs. ≥37 weeks’ gestation). Secondary outcomes were birthweight-for-gestational-age (BW/GA), an indicator of overall fetal growth [50]; percent fat mass (%FM; fat mass divided by total mass), reflecting relative adiposity [51]; and birth length-for-gestational-age (BL/GA), reflecting fetal length accrual and skeletal development [52]. We standardized BW/GA and BL/GA using U.S. natality data [53].

Together, these outcomes capture complementary, biologically related dimensions of fetal growth, skeletal development, and neonatal adiposity.

### Covariates

Using data from medical records, questionnaires, and surveys, we selected covariates for multivariable adjustment based on bivariate associations and *a priori* knowledge. Potential confounders included maternal age, parity, marital status, public assistance use, education, race/ethnicity, geographic region, nativity, pre-pregnancy body mass index (BMI), prenatal smoking, fetal sex, psychiatric diagnosis during pregnancy, and prenatal antidepressant/anxiety medication use. Additionally, we considered gestational diabetes, gestational hypertension, anemia, and preeclampsia as pregnancy complications that affect the birth outcomes but could also be on the causal pathway, with plans to assess their impact in sensitivity analysis.

Additionally, we gave special consideration to maternal race and ethnicity, assessed via self- report surveys using U.S. Census survey categories then consolidated into mutually exclusive categories as Hispanic, non-Hispanic Black, non-Hispanic White, and non-Hispanic Other (including American Indian or Alaska Native, Native Hawaiian or Pacific Islander, multiracial individuals, and those not otherwise specified) to avoid small cell sizes. Race and ethnicity are social constructs that correlate with lived experiences and structural conditions that not only shape both psychosocial stress and birth outcomes but may fundamentally alter the nature of the relationship between psychosocial stress and health outcomes [54–56]. Therefore, we first considered race/ethnicity as an effect modifier, then as a confounder in the absence of evidence for effect modification (i.e., P-interaction below our pre-specified alpha threshold of 0.05).

### Statistical Analyses

Prior to formal analysis, we examined bivariate associations between maternal characteristics and continuous birth outcomes to inform covariate selection.

We conducted the main analysis in three steps. In Step 1, we evaluated associations between the exposure (high vs. low psychosocial distress) and outcomes of interest (gestational age at birth, preterm birth, BW/GA, %FM, and BL/GA) using a series of multivariable linear regression models for continuous outcomes and logistic regression models for dichotomous outcomes. In these models, we first adjusted for maternal age, parity, and pre-pregnancy BMI (Model 1). We tested for effect modification by maternal race/ethnicity using a prenatal distress × race/ethnicity product term. As no evidence of effect modification was observed, Model 2 additionally adjusted for maternal race/ethnicity.

In Step 2, we investigated associations of the potential mediators, maternal ALLO and the ALLO- to-progesterone ratio, with birth outcomes. We assessed the mediators at ∼17 and ∼27 weeks’ gestation, and as change (Δ) between the two timepoints and adjusted for gestational age at blood sample collection as a precision covariate (Model 1), followed by maternal age, parity, and pre-pregnancy BMI (Model 2), and race/ethnicity (Model 3).

In Step 3, we focused on birth outcomes associated with both the exposure and at least one of the mediators. Prior to formal modeling, we tested for interactions between psychosocial distress and each mediator to evaluate for exposure-by-mediator interactions, for which a non-parametric approach to mediation (e.g., the counterfactual-based approach [57]) would be required. Because no significant interactions were observed, we employed the standard regression-based framework [58] to compare the total effect of psychosocial distress on birth outcomes (quantified in Step 1) with the direct effect of psychosocial distress after adjustment for each proposed mediator. We calculated the percent (%) attenuation in the total effect as the percentage reduction in the psychosocial distress regression coefficient after adjustment for each proposed mediator relative to the total effect, with interpreted greater attenuation as more consistent with mediation.

### Sensitivity analyses

We conducted sensitivity analyses adjusting for pregnancy complications, including gestational hypertension, preeclampsia, and anemia, as well as use of antidepressant or anti-anxiety medications.

We ran all models as a complete case analysis, under the assumption of missing completely at random. We defined α = 0.05 as the nominal threshold for statistical significance; however, interpretation of the findings also considered the direction, magnitude, and precision of the effect estimates. We conducted all analyses in R version 4.5.1 (R Core Team, 2025).

## RESULTS

From the original Healthy Start pre-birth cohort of 1,410 individuals, 237 participants met inclusion criteria; 57 were classified as high-distress and 180 as low-distress [18].

Compared with the overall cohort, participants included in this analysis were older (∼1 year), more socioeconomically advantaged as indicated by higher annual household income and educational attainment, and more likely to identify as non-Hispanic White (**Supplemental Table S1**)

Mean ± SD gestational age at birth was 39.47 ± 1.45 weeks. Mean BW/GA z-score was −0.59 ± 0.92, %FM was 8.86 ± 3.99, and BL/GA z-score was −0.18 ± 1.10. Bivariate associations of background characteristics with birth outcomes are presented in **Table 1**. Briefly, gestational length at birth was shorter among participants with pregnancy complications, antidepressants use, and psychiatric diagnoses; BW/GA and BL/GA exhibited similar associations to gestational length at birth with respect to socioeconomic status and pregnancy complications. %FM was inversely associated with parity, prenatal smoking, and male fetal sex; and positively correlated with pre-pregnancy BMI.

**Table 1.**
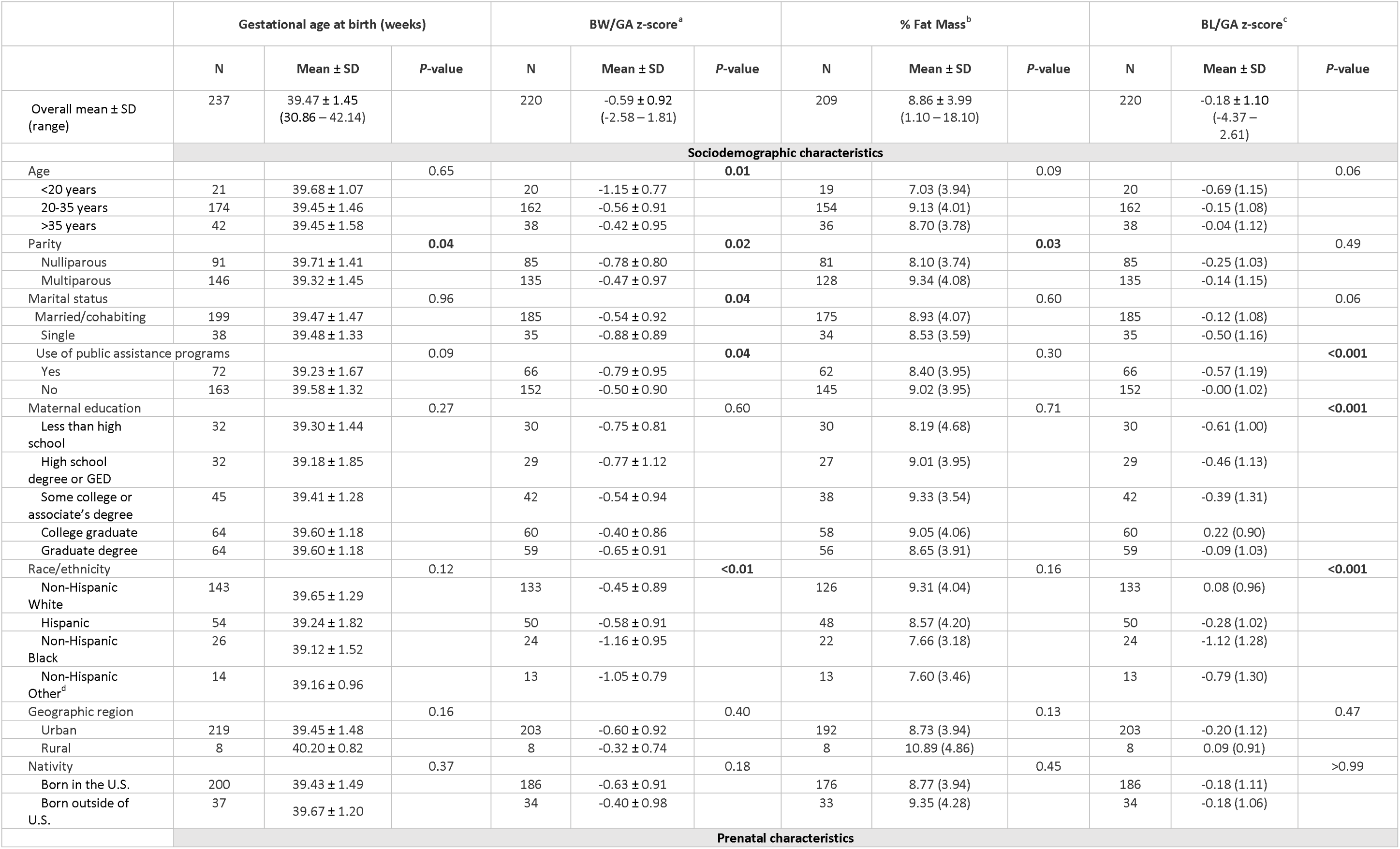
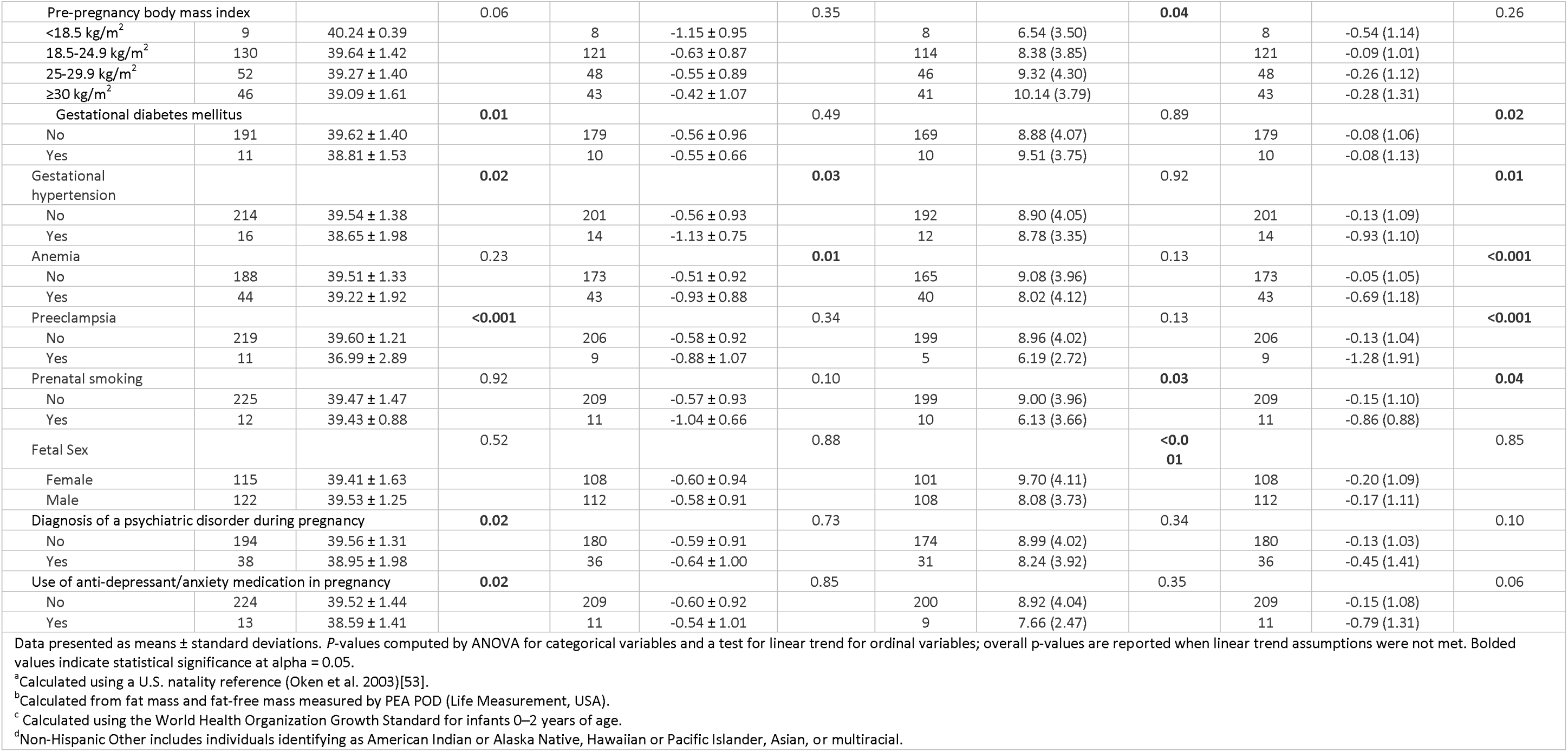
Bivariate associations of maternal sociodemographic and prenatal characteristics with gestational age at birth, birthweight-for-gestational age z-score (BW/GA), percent fat mass fat mass), and birth length-for-gestational age z-score (BL/GA) among pregnant individuals in the Healthy Start Cohort

**Table 2** presents associations between prenatal psychosocial distress and birth outcomes. Psychosocial distress was not associated with gestational age at birth, BW/GA, or %FM in unadjusted and adjusted models, but was associated with shorter BL/GA. In unadjusted models, high vs. low psychosocial distress was associated with a 0.61-unit lower BL/GA z-score (β = −0.61; 95% CI: −0.94, −0.27). This association persisted after adjustment for maternal age, parity, and pre-pregnancy BMI (β = −0.49; 95% CI: −0.84, −0.14), but was attenuated toward the null after adjustment for race/ethnicity (β = −0.35; 95% CI: −0.70, 0.00). Psychosocial distress was not associated with preterm birth, though estimates were imprecise due to the low number of preterm births (n=10). Adjusting for pregnancy complications and medication use did not materially change these findings (**Supplemental Table S2**).

**Table 2.**
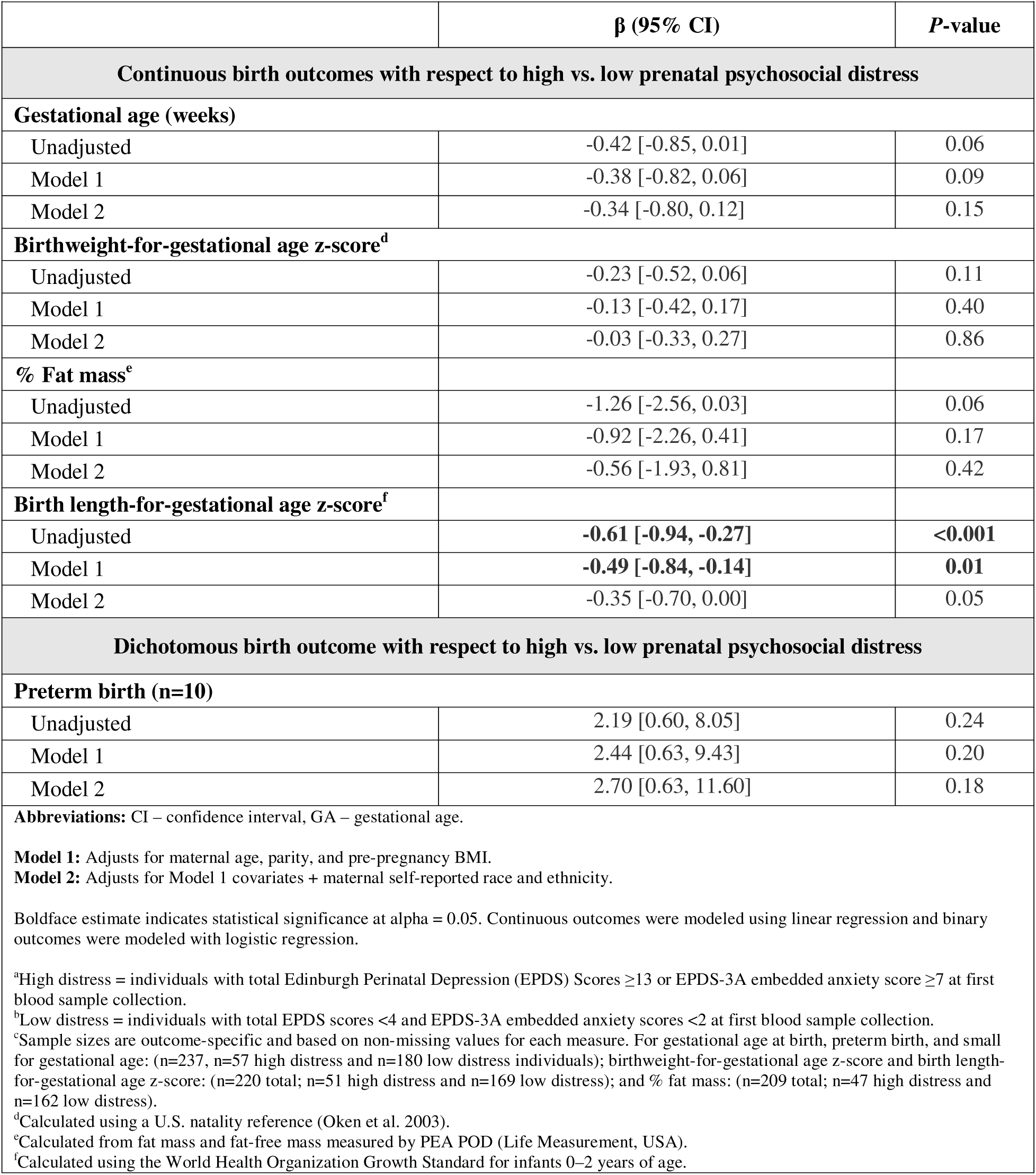
Associations of higha (n = 57) vs. lowb (n=180) maternal psychosocial distress with birth outcomes among 237c pregnant participants in the Healthy Start Cohort.

**Tables 3** and **4** present associations of the potential mediators, maternal serum ALLO and ALLO-to-progesterone ratio, with birth outcomes. Neither variable at either pregnancy time point was consistently associated with gestational age at birth, BW/GA, or %FM. Higher ALLO concentrations at ∼27 weeks’ gestation and a higher ALLO-to-progesterone ratio at both pregnancy time points were associated with greater BL/GA, although only the association for the ∼27-week ALLO-to-progesterone ratio remained after full adjustment. ΔALLO and the ΔALLO-to-progesterone ratio were not associated with any birth outcomes (**Supplemental Tables S3** and **S4**).

**Table 3.**
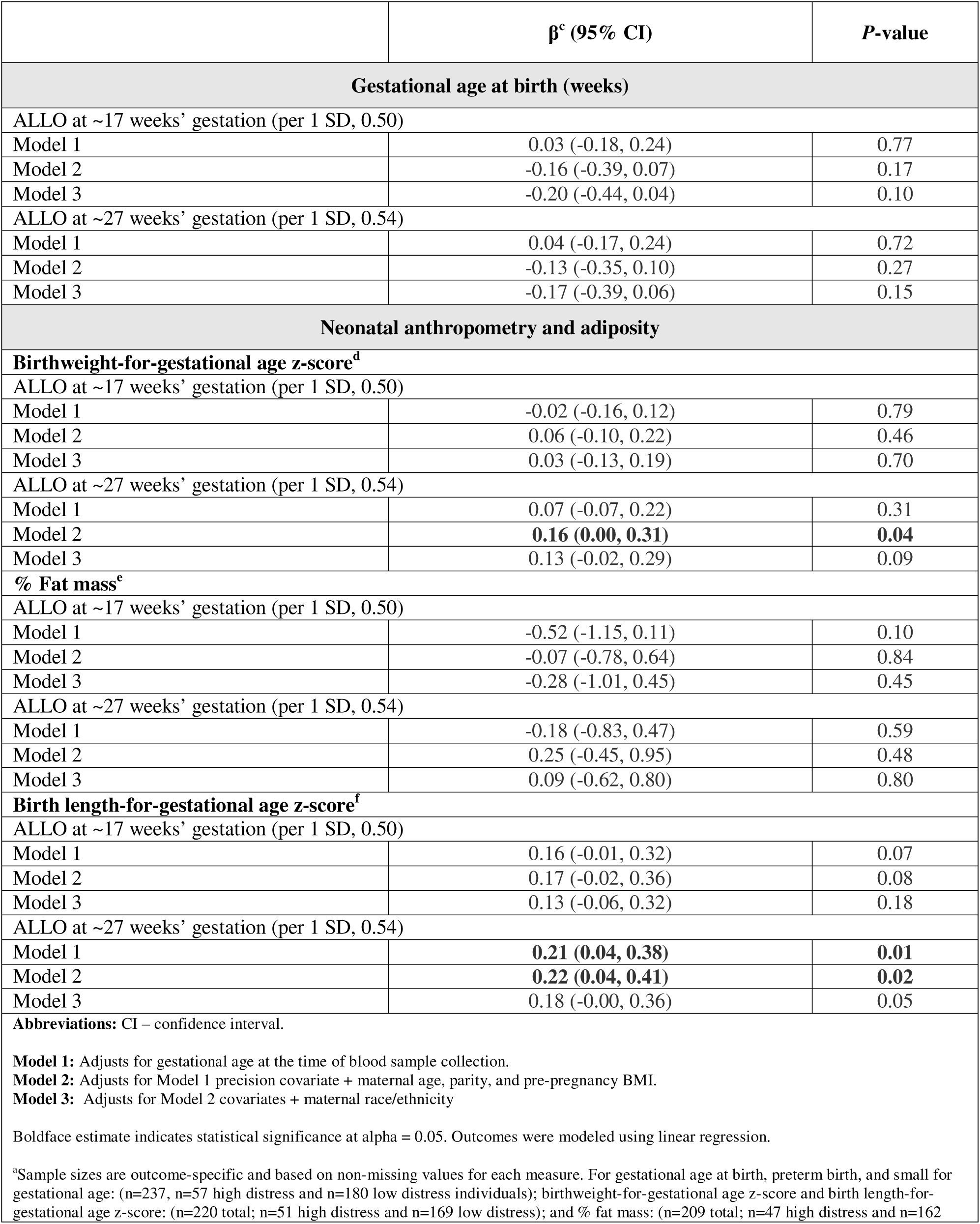
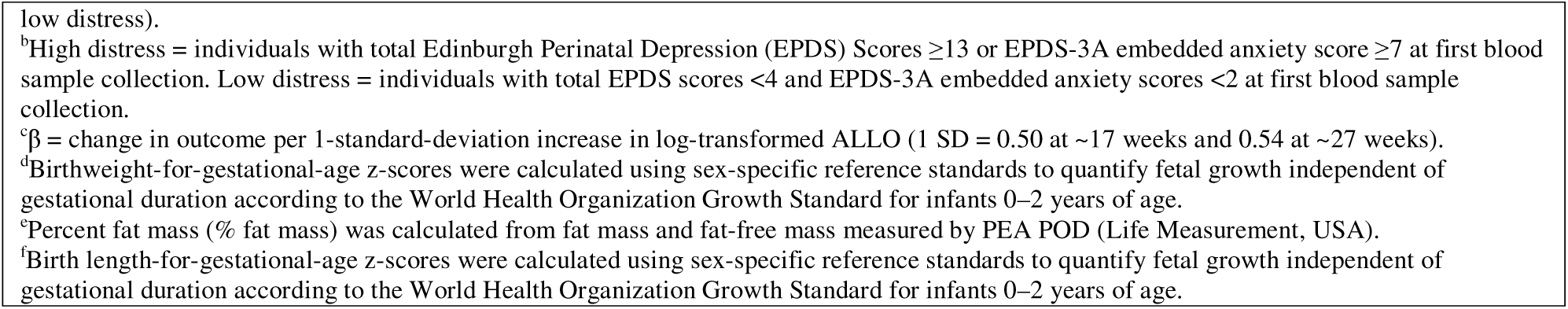
Associations of maternal serum allopregnanolone (ALLO) during early and mid-pregnancy with birth outcomes among 237a pregnant participants in the Healthy Start Cohort enriched for high and low psychosocial distressb

**Table 4.**
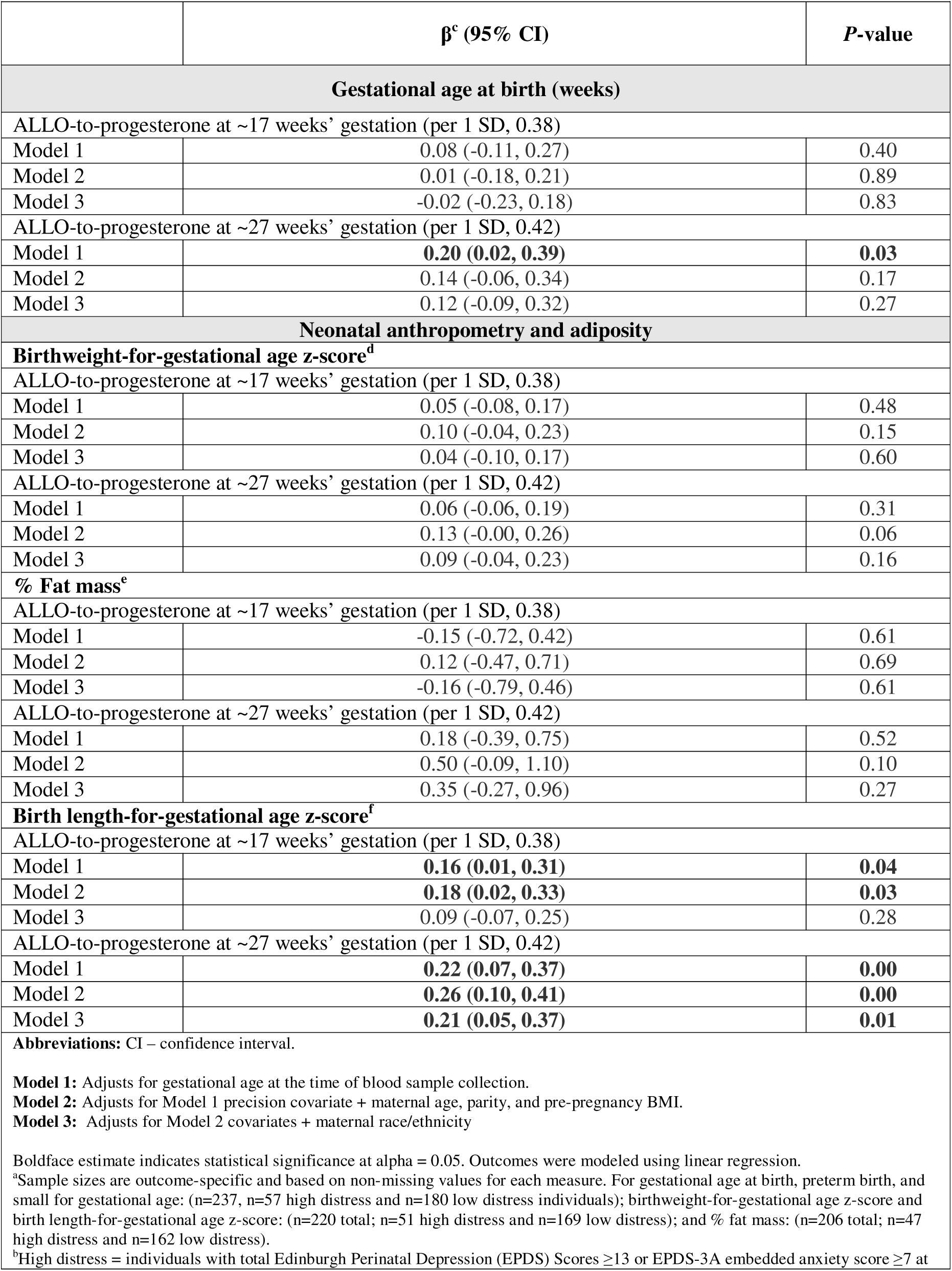
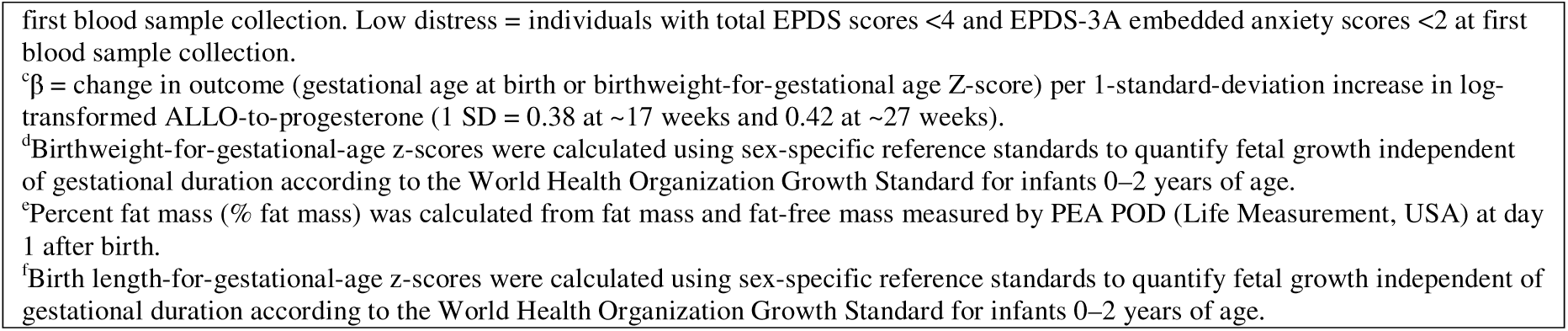
Associations of maternal serum ALLO-to-progesterone ratio during early and mid-pregnancy with birth outcomes among 237a pregnant participants in the Healthy Start Cohort enriched for high and low psychosocial distressb

**Table 5** presents results from the mediation analysis, which took place among 220 of the 237 mother-offspring pairs in the analytic sample for whom we had complete data on the exposure and mediators (no missing values), but not secondary outcomes (n=17 missing values for BL/GA). Here, we examined the relationship between maternal prenatal distress and BL/GA, focusing on ALLO and ALLO-to-progesterone ratio measured at ∼27 weeks’ gestation based on the preceding analytical steps. Adjustment for ALLO resulted in minimal attenuation of the association between prenatal psychosocial distress and BL/GA (β = −0.46; 95% CI: −0.81, −0.10), corresponding to 6.1% attenuation of the total effect. Adjustment for the ALLO-to-progesterone ratio resulted in greater attenuation of the association (β = −0.38; 95% CI: −0.74, −0.03), corresponding to 20.8% attenuation of the total effect, suggesting that the ALLO-to-progesterone ratio may partially mediate the relationship between maternal prenatal distress and shorter BL/GA.

**Table 5.** Mediation analysis of the association between distress status and birth length, with allopregnanolone (ALLO) and the ALLO-to-progesterone ratio as potential mediators among 220 pregnant participants in the Healthy Start Cohort.

|  |  |  |  |
| --- | --- | --- | --- |
| **Mediation Model** | **β [95% CI]** | **P-value** | **% Attenuation** |
| Total effect<sup>a</sup> of maternal distress on birth length<sup>b</sup> | -0.49 [-0.84, -0.14] | 0.01 | — |
| **ALLO** |  |  |  |
| Direct effect<sup>c</sup> of maternal distress on birth length (adjusted for (ln)ALLO at ~27 weeks) | -0.46 [-0.81, -0.10] | 0.01 | 6.1% |
| **ALLO-to-Progesterone ratio** |  |  |  |
| Direct effect<sup>d</sup> of maternal distress on birth length (adjusted for (ln)ALLO-to-progesterone at ~27 weeks) | -0.38 [-0.74, -0.03] | 0.03 | 20.8% |
| **Exposure:** Psychosocial distress status (high vs low; reference = low distress).<br>**Outcome:** Birth length-for-gestational age z-score.<br>**Mediators:** Standardized log-transformed allopregnanolone (ALLO) at ~27 weeks' gestation. Standardized log-transformed ALLO-to-progesterone ratio at ~27 weeks' gestation.<br><sup>a</sup>Adjusts for maternal age, parity, and pre-pregnancy BMI.<br><sup>b</sup>Birth length is birth length-for-gestational age z-score calculated using the World Health Organization Growth Standard for infants 0–2 years of age.<br><sup>c</sup>Covariates include those in (<sup>a</sup>) + ALLO.<br><sup>d</sup>Covariates included those in (<sup>a</sup>) + ALLO-to-progesterone ratio |  |  |  |

## DISCUSSION

In this cohort enriched for the extremes of prenatal psychosocial distress, higher prenatal psychosocial distress was associated with shorter birth length, as indicated by age- and sex-standardized birth length-for-gestational age z-score (BL/GA). Higher maternal ALLO concentrations and, more consistently, a higher ALLO-to-progesterone ratio during mid-pregnancy (∼27 weeks’ gestation) were associated with longer BL/GA. We also found that the ALLO-to-progesterone ratio partially mediated the association between prenatal psychosocial distress and BL/GA, providing evidence that variation in ALLO neurosteroid biosynthesis, rather than absolute ALLO concentrations, may contribute to the biological pathways linking prenatal psychosocial distress with fetal length accrual.

### Prenatal psychosocial distress and birth length

Our findings extend a growing body of literature suggesting that prenatal psychosocial adversity influences fetal length accrual. A previous study of 604 participants in the Healthy Start reported associations between maternal depressive symptoms during pregnancy and shorter birth length [6]. Similarly, a population-based Danish cohort of more than 78,000 pregnancies found that maternal emotional symptoms were associated with shorter birth length after accounting for gestational age and maternal characteristics, as well as with shorter gestation but not with other measures of neonatal size [59]. Interestingly, in the Danish cohort study, whereas maternal emotional symptoms were associated with shorter birth length, perceived life stress was associated with longer birth length in the same cohort, suggesting that different dimensions of prenatal psychosocial adversity may influence fetal growth through distinct biological pathways. In another study following the World Trade Center terrorist attacks, individuals who were pregnant during the time of the attack and lived within two miles of the site gave birth to infants with shorter birth length compared to the reference group living outside the two-mile range [60]. These findings suggest that birth length may represent a sensitive marker of the intrauterine environment and that prenatal distress may influence fetal linear growth even in the absence of detectable effects on gestational age at birth or neonatal adiposity.

Birth length primarily reflects longitudinal skeletal growth accrued across gestation [52] and is generally less sensitive than birth weight or neonatal adiposity to short-term changes in the in-utero environment, especially during the second half of pregnancy [61]. Accordingly, associations of prenatal exposures with shorter birth length may reflect chronic rather than acute environmental and in utero perturbations. Consistent with this, associations of prenatal psychosocial distress, ALLO, and the ALLO-to-progesterone ratio with BL/GA were attenuated after adjustment for maternal race and ethnicity. Because race and ethnicity reflect differences in lived experiences and exposure to chronic social and structural adversity, this attenuation suggests that features of the maternal social environment contribute meaningfully to the relationship between prenatal psychosocial distress and fetal linear growth. These findings further suggest that prenatal psychosocial distress may influence fetal length accrual through maternal physiological responses shaped by the broader social and structural environment [62–64]. This interpretation is consistent with the Stress-Induced Developmental Plasticity framework, which proposes that maternal physiological systems are highly plastic and responsive to maternal social and environmental conditions through evolutionarily conserved pathways [65]. This framework further suggests that interventions promoting respectful, patient-centered prenatal care and supportive social environments may improve maternal well-being, influence maternal physiological responses to prenatal psychosocial distress, and ultimately benefit fetal growth and birth outcomes [8].

### The ALLO-to-progesterone ratio as a partial mediator

The ALLO-to-progesterone ratio, but not ALLO alone, partially mediated the association between prenatal psychosocial distress and birth length, suggesting that endogenous neurosteroid biosynthesis may represent a biological manifestation of psychosocial distress and a potential pathophysiologic pathway linking prenatal distress with impaired fetal growth. ALLO is synthesized from progesterone through a series of enzymatic steps involving 5α-reductase, which converts progesterone to 5α-dihydroprogesterone (5α-DHP), followed by 3α-hydroxysteroid dehydrogenase, which converts 5α- DHP to ALLO [66]. Accordingly, the circulating ALLO-to-progesterone ratio may capture variation in endogenous neurosteroid biosynthesis by integrating information about both precursor availability and enzymatic conversion, potentially explaining why it was a more informative biomarker than ALLO concentrations alone. This notion is supported by studies of perinatal mood disorders reporting altered progesterone metabolism, including higher concentrations of the intermediate metabolite 5α-DHP – the direct precursor to ALLO – among pregnant individuals with depression [67,68], in addition to reports of general perturbations in progesterone and ALLO in pregnant individuals with anxiety [69].

Given the known anti-inflammatory properties of ALLO [36,70], reduced progesterone-to-ALLO conversion may contribute to shorter fetal length accrual through diminished anti-inflammatory signaling. Prenatal psychosocial distress has been associated with elevated inflammation [71], which impairs fetal skeletal growth in experimental models [72–74]. Future mechanistic studies should investigate whether stress-related alterations in progesterone-to-ALLO conversion contribute to impaired fetal skeletal growth through inflammatory pathways and whether the ALLO-to-progesterone ratio better captures these alterations than ALLO concentrations alone.

### Maternal distress, birth size, and neonatal adiposity

In contrast to the established literature linking prenatal psychosocial distress with shorter gestation and lower birth weight (reviewed in [75,76]), we observed no associations of prenatal psychosocial distress, ALLO, or the ALLO-to-progesterone ratio with gestational age, BW/GA, or neonatal adiposity. While our null findings between ALLO and gestational age are consistent with the limited human literature [40,41], the absence of associations across these outcomes does not preclude a need for future studies investigating whether, how, and why prenatal psychosocial distress and neurosteroid biology may influence specific aspects of fetal development.

### Strengths and limitations

This study has several strengths. We analyzed a well-characterized prospective birth cohort with validated surveys for assessment of psychosocial and mental health during the prenatal period, detailed neonatal phenotyping, and fasting maternal serum samples collected at two time points during pregnancy. We quantified ALLO and progesterone using a rigorously validated HPLC–MS/MS assay, which provided high analytical specificity and sensitivity. We selected participants using an extreme-phenotype design that maximized contrast between high and low psychosocial distress. We also evaluated complementary birth outcomes, both ALLO concentrations and the ALLO-to-progesterone ratio, formal mediation, and sensitivity analyses addressing pregnancy complications and psychotropic medication use.

Several limitations warrant consideration. First, the extreme-phenotype design enriched the sample for psychosocial distress and improved internal contrast but may limit generalizability to the broader obstetric population. Second, the modest sample size, particularly for preterm birth, reduced precision of estimates and limited subgroup analyses. Third, ALLO was measured in maternal serum at two time points during pregnancy and may not fully capture longitudinal variation in neurosteroid exposure across gestation or neurosteroid dynamics within placental and fetal compartments. Fourth, we assumed that missing values for BL/GA (n=17) in the mediation analysis were missing completely at random. While a comparison of background characteristics for these 17 participants vs. those who were included in the mediation analysis did not yield any significant differences, we acknowledge the possibility of systematic missingness and related bias. Finally, the observational design prevents causal inference and maintains the possibility of residual confounding.

### Conclusions

Our findings suggest that prenatal psychosocial distress may influence fetal linear growth, in part, through alterations in neurosteroid biosynthesis, with the ALLO-to-progesterone ratio providing a potentially more informative marker of these alterations than absolute ALLO concentrations. These findings support continued investigation of neurosteroid biosynthesis alongside inflammatory, placental, and metabolic pathways to better understand how maternal physiological adaptation to prenatal psychosocial distress influences fetal growth.

## Data Availability

De-identified participant data and analytic code supporting the findings of this study are available from the corresponding author upon reasonable request and with approval from the Healthy Start Study and the Colorado Multiple Institutional Review Board.

## DECLARATIONS

### Ethics approval

All participants provided written informed consent, and the Colorado Multiple Institutional Review Board approved the parent study (Healthy Start) and this secondary analysis (COMIRB #09-0563 and #24-0224).

### Consent for publications

All authors reviewed and approved this work and consent to the publication of this work.

### Availability of data and material

All data and material are available upon request to the corresponding author.

## Acknowledgments

We thank the LEAD Center biorepository staff for biospecimen preparation and the iC42 research specialists for their assistance with the bioassays.

## REFERENCES

1. Schetter, C. D. et al. Anxiety in Pregnancy and Length of Gestation: Findings From the Healthy Babies Before Birth Study. Heal. Psychol. 41, 894–903 (2022).

2. Schetter, C. D. & Tanner, L. Anxiety, depression and stress in pregnancy. Curr. Opin. Psychiatry 25, 141–148 (2012).

3. Ross, K. M. et al. Pregnancy-specific anxiety and gestational length: The mediating role of diurnal cortisol indices. Psychoneuroendocrinology 153, 106114 (2023).

4. Ramos, I. F. et al. Pregnancy anxiety predicts shorter gestation in Latina and non-Latina white women: The role of placental corticotrophin-releasing hormone. Psychoneuroendocrinology 99, 166–173 (2019).

5. Arvanitidou, O. et al. The Impact of Stress and Depression on the Outcome of Human Gestation. Cureus 15, e48700 (2023).

6. Buck, K. E., Dhaliwal, S. K., Dabelea, D. & Perng, W. Association of maternal psychosocial stress with newborn body composition in the Healthy Start study. J. Dev. Orig. Heal. Dis. 14, 576–583 (2023).

7. Zhang, L. et al. Maternal Prenatal Depressive Symptoms and Fetal Growth During the Critical Rapid Growth Stage. *JAMA Netw*. Open 6, e2346018 (2023).

8. Mayne, G., Buckley, A. & Ghidei, L. Understanding and Reducing Persistent Racial Disparities in Preterm Birth: a Model of Stress-Induced Developmental Plasticity. Reprod Sci 29, 2051–2059 (2022).

9. Wadhwa, P. D., Entringer, S., Buss, C. & Lu, M. C. The Contribution of Maternal Stress to Preterm Birth: Issues and Considerations. Clin Perinatol 38, 351–384 (2011).

10. Purdy, R. H., Morrow, A. L., Moore, P. H. & Paul, S. M. Stress-induced elevations of gamma-aminobutyric acid type A receptor-active steroids in the rat brain. Proc National Acad Sci 88, 4553–4557 (1991).

11. Crowley, S. K. et al. Blunted neuroactive steroid and HPA axis responses to stress are associated with reduced sleep quality and negative affect in pregnancy: a pilot study. Psychopharmacology 233, 1299–1310 (2016).

12. Hellgren, C., Åkerud, H., Skalkidou, A., Bäckström, T. & Sundström-Poromaa, I. Low Serum Allopregnanolone Is Associated with Symptoms of Depression in Late Pregnancy. Neuropsychobiology 69, 147–153 (2014).

13. Osborne, L. M. et al. Lower allopregnanolone during pregnancy predicts postpartum depression: An exploratory study. Psychoneuroendocrino 79, 116–121 (2017).

14. Serra, M. et al. Social Isolation Induced Decreases in Both the Abundance of Neuroactive Steroids and GABAA Receptor Function in Rat Brain. J Neurochem 75, 732–740 (2000).

15. Barbaccia, M. L., Serra, M., Purdy, R. H. & Biggio, G. Stress and neuroactive steroids. Int Rev Neurobiol 46, 243–272 (2001).

16. Evans, J., Sun, Y., McGregor, A. & Connor, B. Allopregnanolone regulates neurogenesis and depressive/anxiety-like behaviour in a social isolation rodent model of chronic stress. Neuropharmacology 63, 1315–1326 (2012).

17. Pisu, M. G. et al. Juvenile social isolation affects maternal care in rats: involvement of allopregnanolone. Psychopharmacology 234, 2587–2596 (2017).

18. Mayne, G. B. et al. Associations of Allopregnanolone and Related Steroid Hormones with Prenatal Psychosocial Distress in the Healthy Start Cohort. (2026). doi:10.64898/2026.07.22.26358684

19. Wenzel, E. S. et al. Levels of neuroactive steroids are elevated in those who develop first- onset depression early in pregnancy. Front. Psychiatry 16, 1557560 (2025).

20. Wenzel, E. S. et al. Neuroactive steroids and depression in early pregnancy. Psychoneuroendocrinology 134, 105424 (2021).

21. Björväng, R. D. et al. Mid-pregnancy allopregnanolone levels and trajectories of perinatal depressive symptoms. Psychoneuroendocrinology 164, 107009 (2024).

22. Deligiannidis, K. M., Kroll-Desrosiers, A. R., Tan, Y., Dubuke, M. L. & Shaffer, S. A. Longitudinal proneuroactive and neuroactive steroid profiles in medication-free women with, without and at-risk for perinatal depression: A liquid chromatography-tandem mass spectrometry analysis. Psychoneuroendocrinology 121, 104827 (2020).

23. Hare, M. M., Barber, A., Shaffer, S. A. & Deligiannidis, K. M. Bidirectional associations between perinatal allopregnanolone and depression severity with postpartum gray matter volume in adult women. Acta Psychiatr. Scand. 150, 404–415 (2024).

24. Grötsch, M. K. & Ehlert, U. Allopregnanolone and mood in the peripartum: a longitudinal assessment in healthy women. Front. Behav. Neurosci. 18, 1499416 (2024).

25. Hellgren, C., Comasco, E., Skalkidou, A. & Sundström-Poromaa, I. Allopregnanolone levels and depressive symptoms during pregnancy in relation to single nucleotide polymorphisms in the allopregnanolone synthesis pathway. Horm. Behav. 94, 106–113 (2017).

26. Paoletti, A. M. et al. Observational study on the stability of the psychological status during normal pregnancy and increased blood levels of neuroactive steroids with GABA-A receptor agonist activity. Psychoneuroendocrino 31, 485–492 (2006).

27. Standeven, L. R. et al. Allopregnanolone and depression and anxiety symptoms across the peripartum: an exploratory study. Arch. Women’s Ment. Heal. 25, 521–526 (2022).

28. Murphy, B. E. P., Steinberg, S. I., Hu, F.-Y. & Allison, C. M. Neuroactive Ring A-Reduced Metabolites of Progesterone in Human Plasma during Pregnancy: Elevated Levels of 5α- Dihydroprogesterone in Depressed Patients during the Latter Half of Pregnancy. J Clin Endocrinol Metabolism 86, 5981–5987 (2001).

29. Schoretsanitis, G. et al. Peripartum allopregnanolone blood concentrations and depressive symptoms: a systematic review and individual participant data meta-analysis. Mol. Psychiatry 30, 1–13 (2024).

30. Patterson, R., Balan, I., Morrow, A. L. & Meltzer-Brody, S. Novel neurosteroid therapeutics for post-partum depression: perspectives on clinical trials, program development, active research, and future directions. Neuropsychopharmacology 49, 67–72 (2024).

31. Meltzer-Brody, S. et al. Brexanolone injection in post-partum depression: two multicentre, double-blind, randomised, placebo-controlled, phase 3 trials. Lancet 392, 1058–1070 (2018).

32. Althaus, A. L. et al. Preclinical characterization of zuranolone (SAGE-217), a selective neuroactive steroid GABAA receptor positive allosteric modulator. Neuropharmacology 181, 108333 (2020).

33. Frye, C. A., Hirst, J. J., Brunton, P. J. & Russell, J. A. Neurosteroids for a successful pregnancy. Ann Ny Acad Sci 14, 1–5 (2010).

34. Brunton, P. J., Russell, J. A. & Hirst, J. J. Allopregnanolone in the brain: Protecting pregnancy and birth outcomes. Prog Neurobiol 113, 106–136 (2014).

35. Brunton, P. J. Endogenous opioid signalling in the brain during pregnancy and lactation. Cell Tissue Res 375, 69–83 (2019).

36. Balan, I. et al. Neuroactive Steroids, Toll-like Receptors, and Neuroimmune Regulation: Insights into Their Impact on Neuropsychiatric Disorders. Life 14, 582 (2024).

37. Patchev, V. K., Shoaib, M., Holsboer, F. & Almeida, O. F. X. The neurosteroid tetrahydroprogesterone counteracts corticotropin-releasing hormone-induced anxiety and alters the release and gene expression of corticotropin-releasing hormone in the rat hypothalamus. Neuroscience 62, 265–271 (1994).

38. McLean, M. et al. A placental clock controlling the length of human pregnancy. Nat Med 1, 460–463 (1995).

39. Paris, J. J., Brunton, P. J., Russell, J. A., Walf, A. A. & Frye, C. A. Inhibition of 5α Reductase Activity in Late Pregnancy Decreases Gestational Length and Fecundity and Impairs Object Memory and Central Progestogen Milieu of Juvenile Rat Offspring. J Neuroendocrinol 23, 1079–1090 (2011).

40. Mayne, G. B. et al. A Nested Case-Control Study of Allopregnanolone and Preterm Birth in the Healthy Start Cohort. J Endocr Soc (2022). doi:10.1210/jendso/bvac179

41. Turkmen, S., Bäckström, T., Flodin, Y. K. & Bixo, M. Neurosteroid involvement in threatened preterm labour. Endocrinol Diabetes Metabolism 4, e00216 (2021).

42. Reddy, U. M. et al. Term Pregnancy. Obstet. Gynecol. 117, 1279–1287 (2011).

43. Razaz, N. et al. Time of delivery among low risk women at 37–42 weeks of gestation and risks of stillbirth and infant mortality, and long term neurological morbidity. Paediatr. Périnat. Epidemiology 36, 577–587 (2022).

44. Espel, E. V., Glynn, L. M., Sandman, C. A. & Davis, E. P. Longer Gestation among Children Born Full Term Influences Cognitive and Motor Development. Plos One 9, e113758 (2014).

45. Sherer, M. L. et al. Biological Mechanisms in Pregnant Women With Anxiety (Happy Mother-Healthy Baby Supplement Study): Protocol for a Longitudinal Mixed Methods Observational Study. JMIR Res. Protoc. 12, e43193 (2023).

46. Crume, T. L. et al. Maternal Fuels and Metabolic Measures During Pregnancy and Neonatal Body Composition: The Healthy Start Study. J Clin Endocrinol Metabolism 100, 1672–1680 (2015).

47. Mayne, G., Bloois, E. D., Dabelea, D. & Christians, U. Development and validation of an LC-MS/MS assay for the quantification of allopregnanolone and its progesterone-derived isomers, precursors, and cortisol/cortisone in pregnancy. Anal Bioanal Chem 413, 5427–5438 (2021).

48. M10 Bioanalytical Method Validation and Study Sample Analysis Guidance for Industry. (2022). at <https://www.fda.gov/media/162903/download>

49. Mazahery, H., Hurst, P. R. von, McKinlay, C. J. D., Cormack, B. E. & Conlon, C. A. Air displacement plethysmography (pea pod) in full-term and pre-term infants: a comprehensive review of accuracy, reproducibility, and practical challenges. Matern. Heal., Neonatol. Perinatol. 4, 12 (2018).

50. Bear, L. M. Early identification of infants at risk for developmental disabilities. Pediatr. Clin. North Am. 51, 685–701 (2004).

51. Moore, B. F., Harrall, K. K., Sauder, K. A., Glueck, D. H. & Dabelea, D. Neonatal Adiposity and Childhood Obesity. Pediatrics 146, e20200737 (2020).

52. Dobbing, J. Foetus into man: Physical growth from conception to maturity, 2nd edition, 1989. Early Hum. Dev. 25, 222–223 (1991).

53. Oken, E., Kleinman, K. P., Rich-Edwards, J. & Gillman, M. W. A nearly continuous measure of birth weight for gestational age using a United States national reference. BMC Pediatr. 3, 6 (2003).

54. Pilgrim, A. & Cities, L. Systemic Racism 101: A Visual History of The Impact of Racism in America. (Adams Media, 2022).

55. Brown, T. H. & Homan, P. Structural Racism and Health Stratification: Connecting Theory to Measurement. J. Heal. Soc. Behav. 65, 141–160 (2024).

56. Flanagin, A., Frey, T., Christiansen, S. L. & Committee, A. M. of S. Updated Guidance on the Reporting of Race and Ethnicity in Medical and Science Journals. JAMA 326, 621–627 (2021).

57. Valeri, L. & VanderWeele, T. J. Mediation Analysis Allowing for Exposure–Mediator Interactions and Causal Interpretation: Theoretical Assumptions and Implementation With SAS and SPSS Macros. Psychol. Methods 18, 137–150 (2013).

58. Baron, R. M. & Kenny, D. A. The Moderator-Mediator Variable Distinction in Social Psychological Research: Conceptual, Strategic, and Statistical Considerations. J Pers Soc Psychol 51, 1173–1182 (1986).

59. Tegethoff, M., Greene, N., Olsen, J., Meyer, A. H. & Meinlschmidt, G. Maternal Psychosocial Adversity During Pregnancy Is Associated With Length of Gestation and Offspring Size at Birth: Evidence From a Population-Based Cohort Study. Psychosom. Med. 72, 419–426 (2010).

60. Lederman, S. A. et al. The Effects of the World Trade Center Event on Birth Outcomes among Term Deliveries at Three Lower Manhattan Hospitals. Environ. Heal. Perspect. 112, 1772–1778 (2004).

61. Camerota, M. & Bollen, K. A. Birth Weight, Birth Length, and Gestational Age as Indicators of Favorable Fetal Growth Conditions in a US Sample. PLoS ONE 11, e0153800 (2016).

62. Mayne, G. B. & Ghidei, L. The impact of devaluing Women of Color: stress, reproduction, and justice. Birth 51, 245–252 (2024).

63. Krieger, N. Embodiment: a conceptual glossary for epidemiology. J Epidemiol Commun H 59, 350 (2005).

64. Mayne, G., Buckley, A. & Ghidei, L. Why Causation Matters: Rethinking “Race” as a Risk Factor. Obstet. Gynecol. (2023). doi:10.1097/aog.0000000000005332

65. Mayne, G., et al. Stress-Induced Developmental Plasticity and Spontaneous Preterm Birth: A Justice-Oriented Eco-Evo-Devo Review. Eur. J. Obstet. Gynecol. Reprod. Biol.: X 27, 100409 (2025).

66. Mellon, S. H., Griffin, L. D. & Compagnone, N. A. Biosynthesis and action of neurosteroids. Brain Res Rev 37, 3–12 (2001).

67. Osborne, L. M. et al. Neuroactive steroid biosynthesis during pregnancy predicts future postpartum depression: a role for the 3α and/or 3β-HSD neurosteroidogenic enzymes? Neuropsychopharmacology 50, 904–912 (2025).

68. Evans, S. E. G., Ross, L. E., Sellers, E. M., Purdy, R. H. & Romach, M. K. 3α-reduced neuroactive steroids and their precursors during pregnancy and the postpartum period. Gynecol Endocrinol 21, 268–279 (2009).

69. Etyemez, S. et al. Metabolites of progesterone in pregnancy: Associations with perinatal anxiety. Psychoneuroendocrinology 156, 106327 (2023).

70. Balan, I. et al. Neurosteroid allopregnanolone (3α,5α-THP) inhibits inflammatory signals induced by activated MyD88-dependent toll-like receptors. Transl. Psychiatry 11, 145 (2021).

71. Ravi, M., Bernabe, B. & Michopoulos, V. Stress-Related Mental Health Disorders and Inflammation in Pregnancy: The Current Landscape and the Need for Further Investigation. Front. Psychiatry 13, 868936 (2022).

72. Swolin-Eide, D., Nilsson, C., Holmang, A. & Ohlsson, C. Prenatal Exposure to IL-1& Results in Disturbed Skeletal Growth in Adult Rat Offspring. Pediatr. Res. 55, 598–603 (2004).

73. Cadaret, C. N. et al. Maternal inflammation at midgestation impairs subsequent fetal myoblast function and skeletal muscle growth in rats, resulting in intrauterine growth restriction at term. Transl. Anim. Sci. 3, txz037 (2019).

74. Benedetti, F. D. The Impact of Chronic Inflammation on the Growing Skeleton: Lessons from Interleukin-6 Transgenic Mice. Horm. Res. Paediatr. 72, 26–29 (2009).

75. Bussières, E.-L. et al. Maternal prenatal stress and infant birth weight and gestational age: A meta-analysis of prospective studies. Dev. Rev. 36, 179–199 (2015).

76. Staneva, A., Bogossian, F., Pritchard, M. & Wittkowski, A. The effects of maternal depression, anxiety, and perceived stress during pregnancy on preterm birth: A systematic review. Women Birth 28, 179–193 (2015)

