## Supplemental Material for "Evaluating Allopregnanolone as a Potential Mediator of Prenatal Psychosocial Distress and Birth Outcomes in the Healthy Start Cohort"

**Supplemental Table S1.** Bivariate associations of maternal sociodemographic, prenatal, and neonatal characteristics comparing the participants included in this study (n=237) to those Healthy Start Cohort participants not included (n=1,173).

| | Included | Not Included | *P*-value[a] |
|---|---|---|---|
| | **n = 237 mean ± SD or N (%)** | **n = 1,173 mean ± SD or N (%)** | |
| **Sociodemographic characteristics** | | | |
| Age (years) | 29 ± 6 | 28 ± 6 | **<0.001** |
| Nulliparous | 91 (38) | 744 (67) | **<0.001** |
| Married/cohabiting | 199 (84) | 931 (80) | 0.20 |
| Use of public assistance program | 72 (31) | 348 (39) | **0.02** |
| Maternal education level | | | **0.01** |
| <High school | 32 (14) | 172 (15) | |
| High school degree or GED | 32 (14) | 227 (19) | |
| Some college or associate's degree | 45 (19) | 289 (25) | |
| College graduate | 64 (27) | 245 (21) | |
| Graduate degree | 64 (27) | 240 (20) | |
| Annual household income ≥$70,000 | 107 (54) | 353 (38) | **<0.001** |
| Race/ethnicity | | | **<0.001** |
| Non-Hispanic White | 143 (60) | 281 (43) | |
| Hispanic | 54 (23) | 270 (42) | |
| Non-Hispanic Black | 26 (11) | 60 (9.2) | |
| Non-Hispanic Other[b] | 14 (5.9) | 39 (6.0) | |
| Geographic region | | | **0.02** |
| Urban | 219 (96) | 955 (99) | |
| Rural | 8 (3.5) | 11 (1.1) | |
| Born in US | 200 (84) | 1,008 (86) | 0.50 |
| **Prenatal characteristics** | | | |
| Pre-pregnancy BMI, $kg/m^2$ | 25.4 ± 6 | 25.6 ± 6 | 0.30 |
| Fetus is female | 115 (49) | 535 (48) | 0.90 |
| Gestational diabetes mellitus | 11 (5.0) | 44 (4.2) | 0.90 |
| Gestational hypertension | 16 (7.0) | 73 (6.6) | 0.90 |
| Anemia | 44 (19) | 226 (20) | 0.60 |
| Preeclampsia | 11 (4.8) | 40 (3.6) | 0.40 |
| Smoked during pregnancy | 12 (5.1) | 90 (7.7) | 0.20 |
| Psychiatric disorder | 38 (16) | 204 (18) | 0.50 |
| Use of anti-depressant/anxiety medication during pregnancy | 13 (5.5) | 57 (4.9) | 0.50 |
| **Neonatal Characteristics** | | | |
| Gestational age at birth (weeks) | 39.47 ± 1.45 | 39.16 ± 2.02 | **0.03** |
| Preterm birth (<37 weeks' gestation) | 10 (4.2%) | 79 (6.9%) | 0.12 |

| | | | |
|---|---|---|---|
| Small for gestational age | 43 (18%) | 107 (9.7%) | **<0.001** |
| Birthweight-for-gestational age z-scores[c] | -0.59 ± 0.92 | -0.69 ± 0.90 | 0.13 |
| % Fat mass[d] | 8.9 ± 4.0 | 9.2 ± 3.9 | 0.40 |
| Birth length-for-gestational age z-scores[e] | -0.18 ± 1.10 | -0.41 ± 1.20 | **0.01** |

Data presented as means ± standard deviations or N (%). Bolded values indicate statistical significance at alpha = 0.05.
[a]*P*-values computed by Chi-square or Student T-tests.
[b]Other includes individuals identifying as American Indian or Alaska Native, Hawaiian or Pacific Islander, Asian, or mixed race.
[c]Birthweight-for-gestational-age z-scores were calculated using sex-specific reference standards to quantify fetal growth independent of gestational duration according to the World Health Organization Growth Standard for infants 0–2 years of age.
[d]Percent fat mass (% fat mass) was calculated from fat mass and fat-free mass measured by PEA POD (Life Measurement, USA) at day 1 after birth.
[e]Birth length-for-gestational-age z-scores were calculated using sex-specific reference standards to quantify fetal growth independent of gestational duration according to the World Health Organization Growth Standard for infants 0–2 years of age.

| **Supplemental Table S2.** Associations of high[a] (n = 57) vs. low[b] (n=180) maternal psychosocial distress with birth outcomes among 237[c] pregnant participants in the Healthy Start Cohort with sensitivity analyses | | |
|---|---|---|
| | **β (95% CI)** | ***P*-value** |
| **Continuous birth outcomes with respect to high vs. low prenatal psychosocial distress** | | |
| **Gestational age (weeks)** | | |
| Unadjusted | -0.42 [-0.85, 0.01] | 0.06 |
| Model 1 | -0.38 [-0.82, 0.06] | 0.09 |
| Model 2 | -0.34 [-0.80, 0.12] | 0.15 |
| *Sensitivity Analyses* | | |
| Model 1 + gestational hypertension | -0.37 [-0.82, 0.08] | 0.11 |
| Model 1 + preeclampsia | -0.27 [-0.69, 0.14] | 0.20 |
| Model 1 + anemia | -0.39 [-0.84, 0.06] | 0.09 |
| Model 1 + antidepressant/anti-anxiety medication | -0.28 [-0.73, 0.17] | 0.23 |
| **Birthweight-for-gestational age z-score[d]** | | |
| Unadjusted | -0.23 [-0.52, 0.06] | 0.11 |
| Model 1 | -0.13 [-0.42, 0.17] | 0.40 |
| Model 2 | -0.03 [-0.33, 0.27] | 0.86 |
| *Sensitivity Analyses* | | |
| Model 1 + gestational hypertension | -0.08 [-0.38, 0.22] | 0.60 |
| Model 1 + preeclampsia | -0.10 [-0.41, 0.21] | 0.53 |
| Model 1 + anemia | -0.09 [-0.39, 0.21] | 0.54 |
| Model 1 + antidepressant/anti-anxiety medication | -0.13 [-0.44, 0.17] | 0.38 |
| **% Fat mass[e]** | | |
| Unadjusted | -1.26 [-2.56, 0.03] | 0.06 |
| Model 1 | -0.92 [-2.26, 0.41] | 0.17 |
| Model 2 | -0.56 [-1.93, 0.81] | 0.42 |
| *Sensitivity Analyses* | | |
| Model 1 + gestational hypertension | -0.39 [-1.81, 1.03] | 0.59 |
| Model 1 + preeclampsia | -0.35 [-1.78, 1.07] | 0.63 |
| Model 1 + anemia | -0.53 [-1.93, 0.86] | 0.45 |
| Model 1 + antidepressant/anti-anxiety medication | -0.43 [-1.83, 0.98] | 0.55 |
| **Birth length-for-gestational age z-score[f]** | | |
| Unadjusted | **-0.61 [-0.94, -0.27]** | **<0.001** |
| Model 1 | **-0.49 [-0.84, -0.14]** | **0.01** |
| Model 2 | -0.35 [-0.70, 0.00] | 0.05 |
| *Sensitivity Analyses* | | |
| Model 1 + gestational hypertension | **-0.45 [-0.81, -0.09]** | **0.01** |
| Model 1 + preeclampsia | **-0.42 [-0.78, -0.06]** | **0.02** |
| Model 1 + anemia | **-0.45 [-0.80, -0.10]** | **0.01** |
| Model 1 + antidepressant/anti-anxiety medication | **-0.44 [-0.79, -0.08]** | **0.02** |
| **Dichotomous birth outcome with respect to high vs. low prenatal psychosocial distress** | | |

| | | |
|---|---|---|
| **Preterm birth (n=10)** | | |
| Unadjusted | 2.19 [0.60, 8.05] | 0.24 |
| Model 1 | 2.44 [0.63, 9.43] | 0.20 |
| Model 2 | 2.70 [0.63, 11.60] | 0.18 |
| *Sensitivity Analyses* | | |
| Model 1 + gestational hypertension | 2.81 [0.67, 11.89] | 0.16 |
| Model 1 + preeclampsia | 3.38 [0.66, 17.27] | 0.14 |
| Model 1 + anemia | 2.55 [0.65, 10.05] | 0.18 |
| Model 1 + antidepressant/anti-anxiety medication | 2.36 [0.58, 9.69] | 0.23 |

**Abbreviations:** CI – confidence interval, GA – gestational age.

**Model 1:** Adjusts for maternal age, parity, and pre-pregnancy BMI.
**Model 2:** Adjusts for Model 1 covariates + maternal self-reported race and ethnicity.

Boldface estimate indicates statistical significance at alpha = 0.05. Continuous outcomes were modeled using linear regression and binary outcomes were modeled with logistic regression.

[a]High distress = individuals with total Edinburgh Perinatal Depression (EPDS) Scores ≥13 or EPDS-3A embedded anxiety score ≥7 at first blood sample collection.
[b]Low distress = individuals with total EPDS scores <4 and EPDS-3A embedded anxiety scores <2 at first blood sample collection.
[c]Sample sizes are outcome-specific and based on non-missing values for each measure. For gestational age at birth, preterm birth, and small for gestational age: (n=237, n=57 high distress and n=180 low distress individuals); birthweight-for-gestational age z-score and birth length-for-gestational age z-score: (n=220 total; n=51 high distress and n=169 low distress); and % fat mass: (n=209 total; n=47 high distress and n=162 low distress).
[d]Calculated using a U.S. natality reference (Oken et al. 2003).
[e]Calculated from fat mass and fat-free mass measured by PEA POD (Life Measurement, USA) at day 1 after birth.
[f]Calculated using the World Health Organization Growth Standard for infants 0–2 years of age.

**Supplemental Table S3.** Associations of the change in maternal serum allopregnanolone (ALLO) from early to mid-pregnancy with birth outcomes among 237[a] pregnant participants in the Healthy Start Cohort enriched for high and low psychosocial distress[b]

| | $\beta^c$ (95% CI) | *P*-value |
|---|---|---|
| **Gestational age at birth (weeks)** | | |
| Δ ALLO (per 1 SD, 0.35) | | |
| Model 1 | 0.01 (-0.19, 0.21) | 0.92 |
| Model 2 | 0.02 (-0.18, 0.21) | 0.87 |
| Model 3 | 0.01 (-0.19, 0.21) | 0.92 |
| **Neonatal anthropometry and adiposity** | | |
| **Birthweight-for-gestational age z-score[d]** | | |
| Δ ALLO (per 1 SD, 0.35) | | |
| Model 1 | 0.12 (-0.01, 0.26) | 0.07 |
| Model 2 | 0.12 (-0.01, 0.25) | 0.08 |
| Model 3 | 0.11 (-0.02, 0.24) | 0.10 |
| **% Fat mass[e]** | | |
| Δ ALLO (per 1 SD, 0.35) | | |
| Model 1 | 0.45 (-0.16, 1.05) | 0.15 |
| Model 2 | 0.35 (-0.24, 0.94) | 0.25 |
| Model 3 | 0.36 (-0.23, 0.94) | 0.23 |
| **Birth length z-score[f]** | | |
| Δ ALLO (per 1 SD, 0.35) | | |
| Model 1 | 0.08 (-0.08, 0.24) | 0.34 |
| Model 2 | 0.08 (-0.08, 0.24) | 0.31 |
| Model 3 | 0.07 (-0.08, 0.22) | 0.37 |

**Abbreviations:** CI – confidence interval.

**Model 1:** Adjusts for gestational age at the time of blood sample collection.
**Model 2:** Adjusts for Model 1 precision covariate + maternal age, parity, and pre-pregnancy BMI.
**Model 3:** Adjusts for Model 2 covariates + maternal race/ethnicity

Boldface estimate indicates statistical significance at alpha = 0.05. Outcomes were modeled using linear regression.

[a]Sample sizes are outcome-specific and based on non-missing values for each measure. For gestational age at birth, preterm birth, and small for gestational age: (n=237, n=57 high distress and n=180 low distress individuals); birthweight-for-gestational age z-score and birth length-for-gestational age z-score: (n=220 total; n=51 high distress and n=169 low distress); and % fat mass: (n=209 total; n=47 high distress and n=162 low distress).
[b]High distress = individuals with total Edinburgh Perinatal Depression (EPDS) Scores ≥13 or EPDS-3A embedded anxiety score ≥7 at first blood sample collection. Low distress = individuals with total EPDS scores <4 and EPDS-3A embedded anxiety scores <2 at first blood sample collection.
[c]β = change in outcome per 1-standard-deviation increase in log-transformed and standardized ALLO (1 SD = 0.35 for delta/change in ALLO).
[d]Birthweight-for-gestational-age z-scores were calculated using sex-specific reference standards to quantify fetal growth independent of gestational duration according to the World Health Organization Growth Standard for infants 0–2 years of age.
[e]Percent fat mass (% fat mass) was calculated from fat mass and fat-free mass measured by PEA POD (Life Measurement, USA).
[f]Birth length-for-gestational-age z-scores were calculated using sex-specific reference standards to quantify fetal growth independent of gestational duration according to the World Health Organization Growth Standard for infants 0–2 years of age.

**Supplemental Table S4.** Associations of the change in maternal serum ALLO-to-progesterone ratios from early to mid-pregnancy with birth outcomes among 237[a] pregnant participants in the Healthy Start Cohort enriched for high and low psychosocial distress[b]

| | **β[c] (95% CI)** | ***P*-value** |
|---|---|---|
| **Gestational age at birth (weeks)** | | |
| Δ ALLO-to-progesterone (per 1 SD, 0.32) | | |
| Model 1 | 0.17 (-0.02, 0.35) | 0.08 |
| Model 2 | 0.14 (-0.04, 0.32) | 0.14 |
| Model 3 | 0.14 (-0.04, 0.32) | 0.14 |
| **Neonatal anthropometry and adiposity** | | |
| **Birthweight-for-gestational age z-score[d]** | | |
| Δ ALLO-to-progesterone (per 1 SD, 0.32) | | |
| Model 1 | 0.03 (-0.10, 0.15) | 0.66 |
| Model 2 | 0.04 (-0.08, 0.16) | 0.50 |
| Model 3 | 0.06 (-0.06, 0.18) | 0.31 |
| **% Fat mass[e]** | | |
| Δ ALLO-to-progesterone (per 1 SD, 0.32) | | |
| Model 1 | 0.38 (-0.18, 0.93) | 0.19 |
| Model 2 | 0.40 (-0.14, 0.94) | 0.15 |
| Model 3 | 0.47 (-0.07, 1.02) | 0.09 |
| **Birth length-for-gestational age z-score[f]** | | |
| Δ ALLO-to-progesterone (per 1 SD, 0.32) | | |
| Model 1 | 0.11 (-0.04, 0.25) | 0.16 |
| Model 2 | 0.11 (-0.04, 0.25) | 0.15 |
| Model 3 | 0.14 (-0.00, 0.28) | 0.05 |

**Abbreviations:** CI – confidence interval.

**Model 1:** Adjusts for gestational age at the time of blood sample collection.
**Model 2:** Adjusts for Model 1 precision covariate + maternal age, parity, and pre-pregnancy BMI.
**Model 3:** Adjusts for Model 2 covariates + maternal race/ethnicity

Boldface estimate indicates statistical significance at alpha = 0.05. Outcomes were modeled using linear regression.

[a]Sample sizes are outcome-specific and based on non-missing values for each measure. For gestational age at birth, preterm birth, and small for gestational age: (n=237, n=57 high distress and n=180 low distress individuals); birthweight-for-gestational age z-score and birth length-for-gestational age z-score: (n=220 total; n=51 high distress and n=169 low distress); and % fat mass: (n=206 total; n=47 high distress and n=162 low distress).

[b]High distress = individuals with total Edinburgh Perinatal Depression (EPDS) Scores ≥13 or EPDS-3A embedded anxiety score ≥7 at first blood sample collection. Low distress = individuals with total EPDS scores <4 and EPDS-3A embedded anxiety scores <2 at first blood sample collection.

[c]β = change in outcome (gestational age at birth or birthweight-for-gestational age Z-score) per 1-standard-deviation increase in log ALLO-to-progesterone (1 SD = 0.32 for delta/change).

[d]Birthweight-for-gestational-age z-scores were calculated using sex-specific reference standards to quantify fetal growth independent of gestational duration according to the World Health Organization Growth Standard for infants 0–2 years of age.

[e]Percent fat mass (% fat mass) was calculated from fat mass and fat-free mass measured by PEA POD (Life Measurement, USA) at day 1 after birth.

[f]Birth length-for-gestational-age z-scores were calculated using sex-specific reference standards to quantify fetal growth independent of gestational duration according to the World Health Organization Growth Standard for infants 0–2 years of age.
